# Automated Classification of Fat-infiltrated Axillary Lymph Nodes on Screening Mammograms

**DOI:** 10.1101/2022.09.20.22280111

**Authors:** Qingyuan Song, Roberta M. diFlorio-Alexander, Ryan T. Sieberg, Dennis Dwan, William Boyce, Kyle Stumetz, Sohum D. Patel, Margaret R. Karagas, Todd A. Mackenzie, Saeed Hassanpour

## Abstract

**Objectives:** Fat-infiltrated axillary lymph nodes (LNs) are unique sites for ectopic fat deposition. Early studies showed a strong correlation between fatty LNs and obesity-related diseases. Large-scale studies are needed to confirm these preliminary results but this is hampered by the scarcity of labeled data. With the long-term goal of developing a rapid and generalizable tool to aid data labeling, we developed an automated deep learning (DL)-based pipeline to classify the status of fatty LNs on screening mammograms.

**Methods:** Our internal dataset included 886 mammograms from a tertiary academic medical institution, with a binary status of the fat-infiltrated LNs based on the size and morphology of the largest visible axillary LN. A two-stage DL model training and fine-tuning pipeline was developed to classify the fat-infiltrated LN status using the internal training and development dataset. The model was evaluated on a held-out internal test set and a subset of the Digital Database for Screening Mammography.

**Results:** Our model achieved an accuracy of 0.97 (95% CI: 0.94-0.99) on 264 internal testing mammograms and an accuracy of 0.82 (95% CI: 0.77-0.86) on 70 external testing mammograms. The model successfully extracted meaningful LN-related features from the mammograms.

**Conclusion:** This study confirmed the feasibility of using a DL model for fat-infiltrated LN classification. The model provides a practical tool to identify fatty LNs on mammograms and to allow for future large-scale studies to evaluate the role of fatty LNs as an imaging biomarker of obesity-associated pathologies.

**Advances in knowledge:** Our study is the first to classify fatty LNs using an automated DL approach.

## Introduction

Obesity is highly prevalent worldwide and is a substantial risk factor for increased morbidity and mortality from cardiovascular disease, cancer and other chronic diseases^1–3^. An increasing body of evidence shows that body mass index (BMI) > ^30 kg/m2^ is insufficient for determining the risk of obesity-related diseases^4–6^. In contrast, ectopic fat deposition within and around organs such as the visceral cavity, liver, and muscle is a better predictor of obesity-associated illness and can be quantified from medical imaging such as MRI and CT ^4,7,8^. Ongoing work suggests that axillary lymph nodes (LNs) are novel ectopic fatty depots associated with obesity-related disease^9,10^. However, annotated data in this domain are limited, and further large-scale studies are limited by the lack of an automated tool to annotate and characterize fat-infiltrated LNs.

Benign, nonmetastatic LNs demonstrate variable size and morphology due to variable hilar fat deposition. On mammography, fat-infiltrated LNs have an expanded, lucent, fat-infiltrated hilum that enlarges the overall size of the LN, while normal axillary LNs are smaller and radiologically denser (**Figure 1**). Recent research has shown that fat-infiltrated axillary LNs identified on screening mammograms are highly linked to obesity^11^. A recent case□control study of 431 obese breast cancer patients further showed that patients with enlarged fat-infiltrated contralateral axillary LNs on breast MRI and mammography had a higher risk of ipsilateral breast cancer nodal metastases, independent of patients’ age, BMI, and tumor characteristics^9^. Another recent study reported a positive association between fat-infiltrated axillary LNs and the prevalence of type 2 diabetes (T2DM) in women with obesity, independent of age and BMI, and confirmed that fat-infiltrated LNs are more commonly seen in women with obesity^10^. A smaller study recently reported a decrease in serum lipids in obese women who had a reduction in the size of fatty nodes after bariatric surgery, raising the possibility that fatty LNs are a modifiable risk factor for cardiometabolic disease among people with obesity^12^. These studies highlight the potential clinical importance of mammographically detected fat-infiltrated axillary LNs as a novel imaging biomarker of an advanced breast cancer status and cardiometabolic disease among women with obesity.

**Figure 1.**
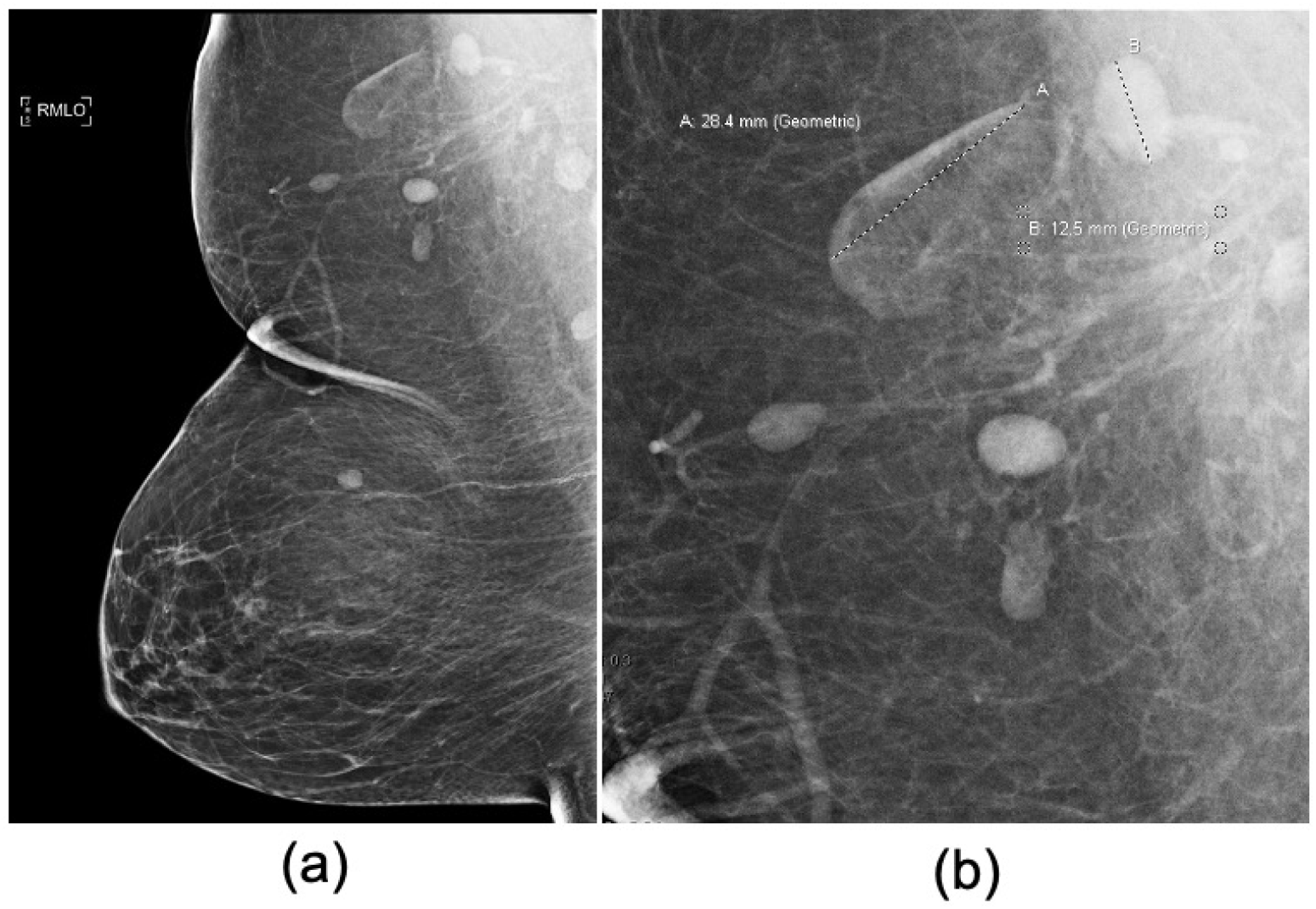
Screening mammogram of a female (70’s) with benign variable LN morphology. **a)** Full-field mediolateral oblique (MLO) view of the right breast and **b)** magnified view of the right axilla demonstrates two adjacent LNs with differences in central lucency and size and morphology (A: 28 mm versus B: 3 mm) secondary to variable degrees of fat expansion of the LN hilum. We recorded the status as a benign fat-infiltrated LN based on the largest longitudinal axis of the largest visible axillary LN.

Axillary LNs are visualized on approximately 50%-80% of digital screening mammograms^11,13^. Standard clinical mammographic imaging interpretation includes the evaluation of the axilla for abnormal LNs that may indicate a malignant process. However, fat-infiltrated LNs are considered benign normal variants that do not require further evaluation, measurement, or analysis. In the studies mentioned above, the measurement and classification of axillary LNs were obtained manually and confirmed by multiple radiology experts^9,11,14^. The manual assessment procedure is not only time-consuming and laborious but can also result in interobserver inconsistencies. Previous studies have reported variable interrater agreement for the classification of axillary LNs, with Pearson correlation coefficients ranging from 0.64 to 0.95, suggesting that the reader agreement can be improved^9,11,12^. An automated pipeline is required to standardize the labeling process and speed up the assessment of the fat-infiltrated LN status on breast images in bulk for future large-scale studies to further explore the potential of fat-infiltrated LNs as imaging biomarkers of obesity associated diseases.

In recent years, deep learning (DL) models such as convolutional neural networks (CNNs) have been utilized for medical image analysis and demonstrated their usefulness in various clinical tasks^15–17^. CNN approaches can bypass the laborious manual feature extraction steps and directly operate on the input images by learning the indicative patterns for the classification and characterization of the images.

Additionally, CNN models that have been pretrained on vast image datasets and have learned feature extraction on generic images can be subsequently fine-tuned to attain expertise-level performance on medical image datasets that are usually much smaller^18,19^.

In this paper, we aim to investigate the feasibility of employing DL models for fat-infiltrated LN assessments on screening mammograms, with the long-term goal of establishing an automated, end-to-end fat-infiltrated LN classifier for future studies. The purpose of this study is to lay the groundwork for future research investigating fat-infiltrated LNs as potential imaging biomarkers of obesity-related pathologies. To the best of our knowledge, this is the first attempt to use a DL technique to automatically evaluate the status of fat-infiltrated LNs on screening mammography.

## Methods

### Data Source for Model Development

This study, and the usage of human participant data in this project, were approved by the Institutional Review Board (IRB) with a waiver of informed consent. The internal dataset used for this study included 886 full-field digital screening mammogram (FFDM) mediolateral oblique (MLO) views with detectable axillary LNs collected through a retrospective review of screening mammograms over an 8-month period at a tertiary care academic medical facility. The detailed data collection process is described in a previous study^10^. In brief, the largest visible axillary LN on the left or right MLO view was measured and labeled, with a final assessment achieved via a consensus review between two radiology reviewers^9^. The images were labeled as positive samples if they contained a fat-infiltrated LN larger than 18 mm and with a fat-infiltrated LN morphology, or as negative samples if smaller than or equal to 18mm. The dataset yielded 416 positive samples and 470 negative samples.

### Image Preprocessing and Internal Dataset Construction

The mammograms were received in the standard Digital Imaging and Communications in Medicine (DICOM) format and came in various orientations and sizes. Images were converted to JPEG format before applying the preprocessing workflow implemented in Python (version 3.8). We first identified and applied a binary mask using the largest contour of the breast and removed all text markers and noise outside of the contour of the breast. Through visual inspection of the mammograms, we found that a region of 1500×1500 pixels in the axillary region of the mammogram was sufficient to cover all the axillary LNs. Therefore, a 1500×1500-pixel patch in the axillary region of each image was extracted and used to construct the initial dataset for model development.

### Oversampling of Nonaxillary Patches

The axillary region of mammograms contains radiographic densities that are unrelated to the imaging characteristics of the axillary LNs. These densities, including but not limited to breast tissue, vessels, muscle, skin folds, fibro-glandular tissue and radiographic densities that are also present in the non-axillary portion of the mammogram. To help the model better distinguish the LNs from non-nodal radiographic densities, we sampled patches from nonaxillary regions of the breast and used them as additional negative samples. We implemented a sliding-window approach to sample additional 1500×1500 pixel images outside the axillary region, with a step size of 750 pixels, as illustrated in **Figure** 2. These additional nonaxillary patches were combined into the negative samples of the initial dataset.

**Figure 2.**
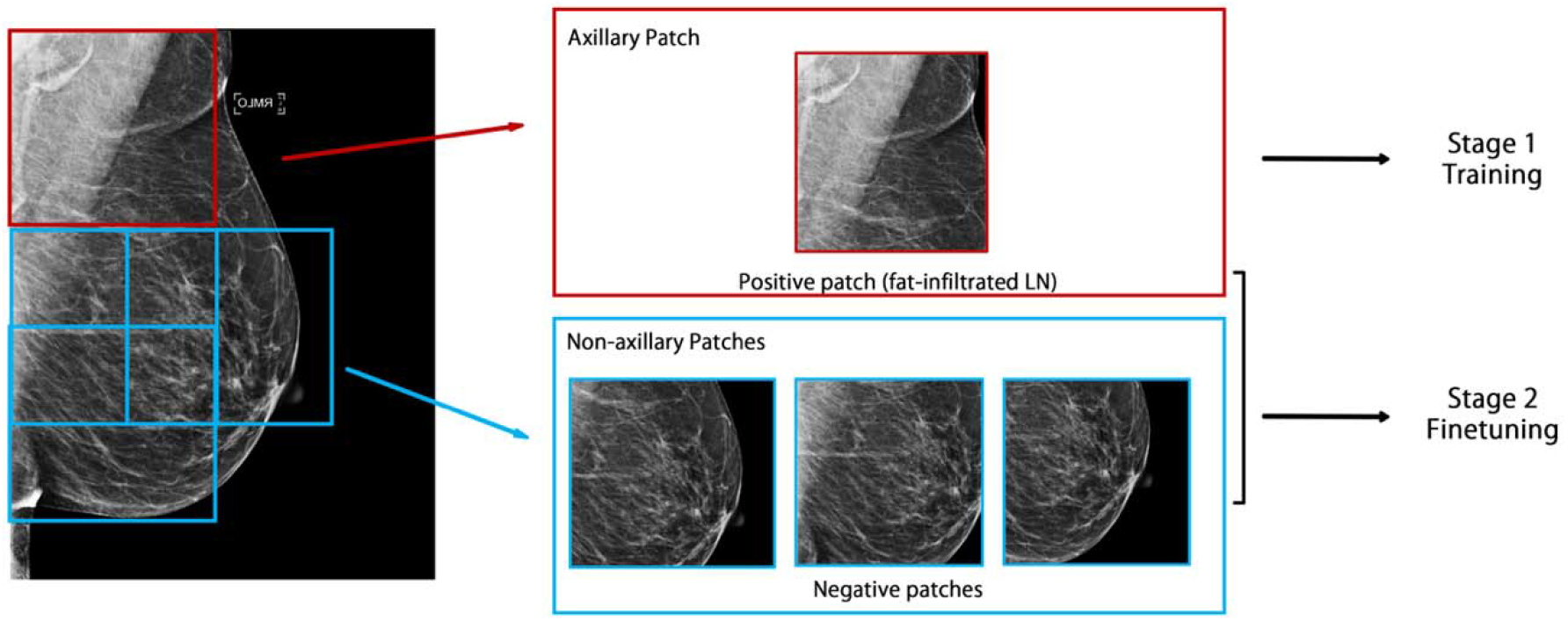
Automated image processing and dataset construction. A patch was extracted from the upper outer corner of the MLO view and used as the axillary patch during the model training and hyperparameter tuning stage. Additional patches were extracted by a sliding window approach from the nonaxillary region and added to the negative samples in the model-finetuning stage.

Finally, we randomly split the dataset into a 70% training set, a 10% development set for hyperparameter tuning, and a 20% testing set. **Table 1** shows the class distribution of the initial dataset’s training, development and testing sets, and the oversampled larger dataset that included the nonaxillary patches.

**Table 1.**
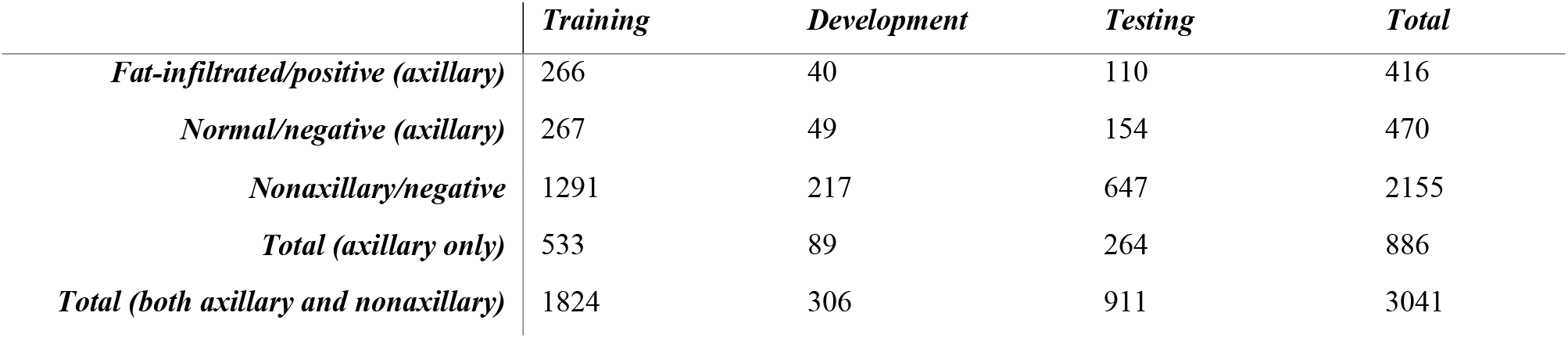
The distribution of mammograms in the training, development and test sets.

|  | *Training* | *Development* | *Testing* | *Total* |
| --- | --- | --- | --- | --- |
| *Fat-infiltrated/positive (axillary)* | 266 | 40 | 110 | 416 |
| *Normal/negative (axillary)* | 267 | 49 | 154 | 470 |
| *Nonaxillary/negative* | 1291 | 217 | 647 | 2155 |
| *Total (axillary only)* | 533 | 89 | 264 | 886 |
| *Total (both axillary and nonaxillary)* | 1824 | 306 | 911 | 3041 |

### Model Training

The model training was conducted in two stages, a model training and selection stage, followed by a model-finetuning stage. In the first stage, various CNN models were trained on the smaller dataset with only axillary patches to select the best-performing model architecture and hyperparameter settings. We experimented with several well-established CNN architectures, including VGG16^20^, ResNet18^21^ and DenseNet121^22^, with initial weights pretrained on the ImageNet dataset^23^. Kaiming weight initialization^24^was used for the binary output fully connected layer. Hyperparameters, including batch size (8 or 16) and initial learning rate (1 × 10^−4^, 5 × 10^−5^, or 2.5 × 10^−5^), were selected by a grid search to optimize the cross-entropy loss. The model with the lowest loss on the development set was selected for the fine-tuning stage. In the second stage, the pretrained model chosen from the previous stage was further fine-tuned on a larger training dataset that included the oversampled negative patches using the best hyperparameters selected from the first stage. In both stages, the model parameters were optimized using the ADAM optimizer^25^ with a step learning rate decay of 0.5 every 15 epochs throughout the training. The training ended when the development set loss did not show improvement for 30 consecutive epochs.

Before training, grayscale image patches were resized to 224×224 pixels and converted to 3-channel images by repeating it three times to fit into the pretrained model that operates on RGB images. Images were standardized to have a mean of zero and a standard deviation of one. To expand the training data’s size and improve the model’s generalizability, we used data augmentation on the training data, including random 90-degree rotation, random horizontal and vertical flipping, sharpening, noise addition, and contrast variation. The details of data preprocessing and augmentation implementation are described in **Supplementary Material S1**. The model training process was conducted on NVIDIA Titan Xp graphical processing units (GPUs) with 12 GB of memory and was implemented using the PyTorch framework^26^.

### Internal Model Evaluation and Visualization

We evaluated the performance of the final model on the test set by calculating the accuracy, sensitivity, specificity, precision, sensitivity and F1 score. Confidence intervals were calculated with 10,000 bootstrap resampling. We also plotted the receiver operating characteristic (ROC) curve and calculated the area under the curve (AUC). To address the interpretability of our DL model, we generated the visualization of our model outcome on the images using Gradient-weighted Class Activation Mapping (GradCAM)^27^. The resulting heatmaps highlighted the most indicative region of the images based on the region’s influence on the model’s prediction outcome.

### External Validation

To assess the generalizability of our model, we evaluated it on images from the Curated Breast Imaging Subset of the Digital Database for Screening Mammography (CBIS-DDSM). This dataset contains scanned film mammography studies in DICOM format. From the dataset, 110 normal mammograms without any abnormalities (Breast Imaging Reporting and Data System (BI-RADS) category 1) and with visible axillary LNs were sampled as the external test set for this study. A breast radiologist, and a graduate student with four years of experience in FIN radiology, independently reviewed the cases and labeled them as fat-infiltrated or normal. Due to the loss of pixel spacing information in this dataset, the size of the LN could not be measured directly from the mammograms. Therefore, an LN was labeled fat-infiltrated if it displayed a fat-infiltrated LN morphology with an expanded fatty hilum relative to the surrounding nodal cortex as previously described. Cases with disagreement were further discussed, and a final classification was determined by consensus between the two readers. Cases with low visibility due to multiple overlapping nodes were removed from this external test set. The review yielded 35 cases with fat-infiltrated LNs, and 35 cases with normal axillary nodes, to construct a balanced dataset. We evaluated our model on the labeled external data and visualized the model decision with GradCAM.

## Results

### Model Performance on the Internal Dataset

During stage 1 of training, the ResNet18 model outperformed the other two architectures with a testing accuracy of 0.94 with 15 false predictions out of 264 testing samples (**Table 2a**). Therefore, the ResNet18 model was chosen for further fine-tuning. The model performance was improved by fine-tuning the model on the oversampled negative patches from the nonaxillary region with a testing accuracy of 0.97 on the axillary patches. The final hyperparameters for each model, based on our parameter tuning on the development set, are shown in **Supplementary Table 1**, and the comparison of the Stage 1 model performance is included in **Supplementary Figure 1**. The visualization of the model’s outcome highlighted by the GradCAM heatmap (**Figure 3**) indicated that the model considered the contributing features of both fat-infiltrated and normal LNs in its decision-making. In addition, the model correctly classified the cases as fat-infiltrated, when both the fat-infiltrated and normal LNs were present on the mammograms, by focusing on only the regions of the fat-infiltrated node **(Figure 3b)**.

**Figure 3.**
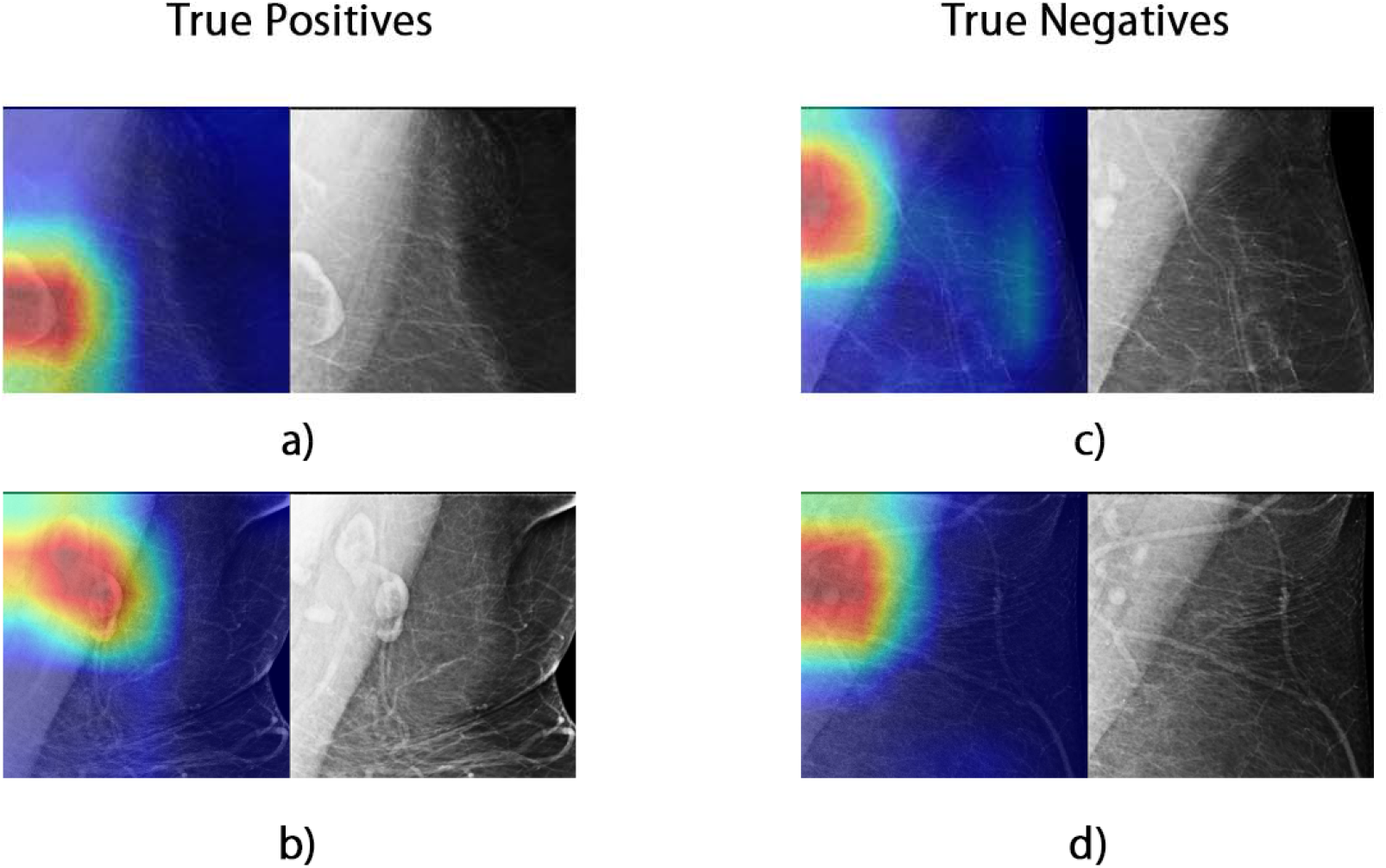
Examples of Grad-CAM visualization. Red highlights the areas that highly influenced the model’s outcome, while blue indicates regions considered unimportant to the predictions by the model. The original grayscale images are shown on the side for reference.

**Table 2.**
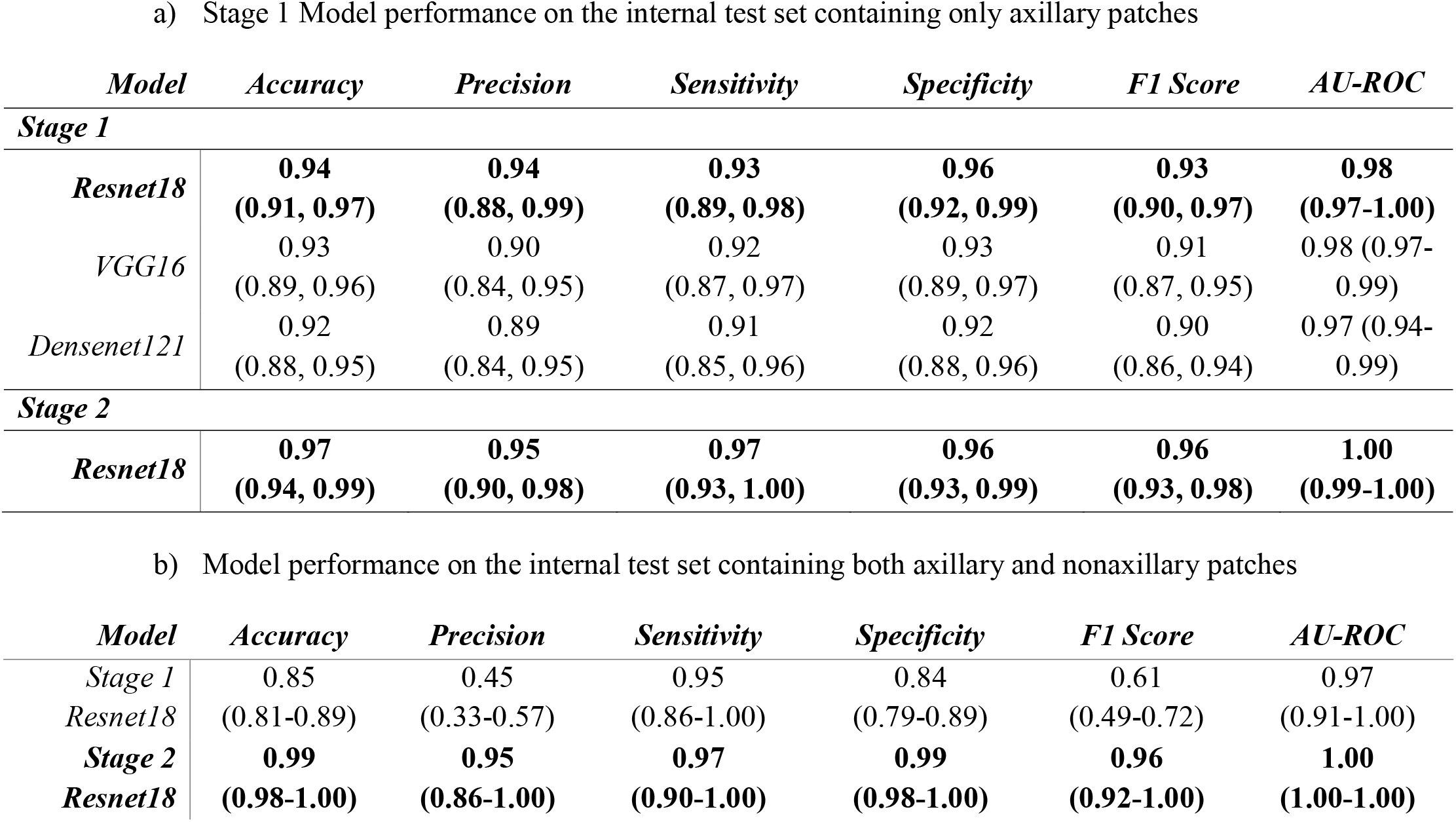
Summary of the Evaluation Metrics of Best-performed Models on the Internal Test Set with 95% Confidence Intervals.

Of note, although the best-performing model from Stage 1 of training achieved a satisfactory classification performance on the axillary patches, the model was prone to make false predictions by misclassifying features unrelated to fat-infiltrated LNs. Through visual inspection and analysis of the model errors, we found that false-positives were caused by misinterpretation of an adjacent dense vessel as an LN cortex or mischaracterization of small normal LNs grouped around central subcutaneous fat as a single larger node with expanded hilar fat. The false negative classifications were due to the model failing to localize the LN due to dense muscle or breast tissue, or when the model focused only on the normal LNs when a mixture of normal and fat-infiltrated LNs was present. An example visualization of the errors is shown in **Supplementary Figure 2**. The errors from the Stage 1 Resnet18 model were more prominent when applying the model to the oversampled test set (**Table 2b**). The model yielded more false-positive classifications by mistakenly classifying breast tissue and vessels as fat-infiltrated LNs. After fine-tuning the model with oversampled negative patches, the errors were significantly reduced (**Table 2b**), and the AU-ROC was improved to 1.00 (**Figure 4**).

**Figure 4.**
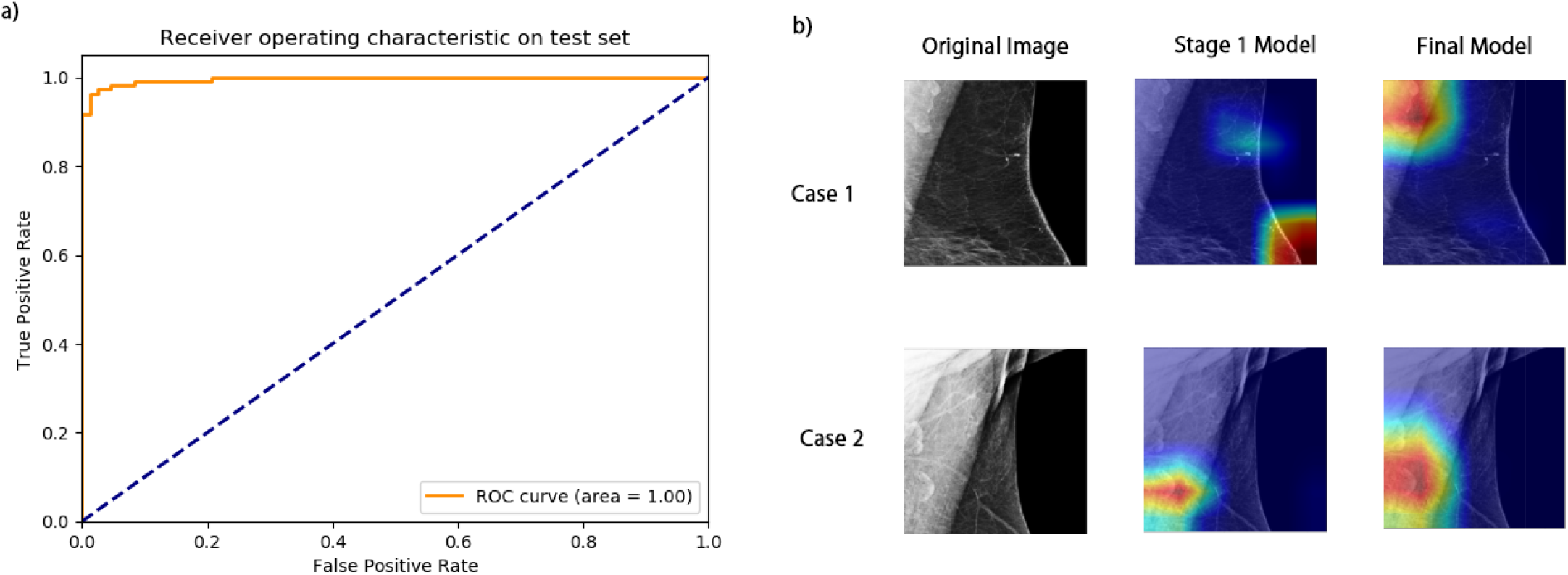
Stage 2 model AU-ROC and examples of improved model predictions. a) The AU-ROC of the Stage 2 model improved to 1.00. b) Two cases illustrate the improved final model performance. In Case 1, the final model correctly localized the axillary region, which the Stage 1 model failed to. In Case 2, the final model more accurately highlighted the entire fat-infiltrated LN, while the Stage 1 model only highlighted part of the node.

### Model Performance on External Validation

On the external dataset, the Pearson correlation between the independent labels of the two reviewers was 0.61 (p value 0.001). Using the consensus labels of the external data, our model achieved an accuracy of 0.82 (95% CI: 0.77-0.86) and an AUROC of 0.87 (95% CI: 0.82-0.91) (**Table 3**), and the visualization of model performance on the external data is shown in **Figure 5**. Overall, the model identified most of the cases with fat-infiltrated nodes (sensitivity: 0.92, 95% CI: 0.87-0.96). False negatives resulted from failing to identify the fat-infiltrated LN from the background muscle due to low contrast (**Figure 5c**). The model resulted in more false-positives on the external dataset than on the internal dataset. The false-positives were mostly due to misclassifying a cluster of multiple normal nodes or surrounding vessels as fat-infiltrated nodes (**Figure 5d-f**).

**Figure 5.**
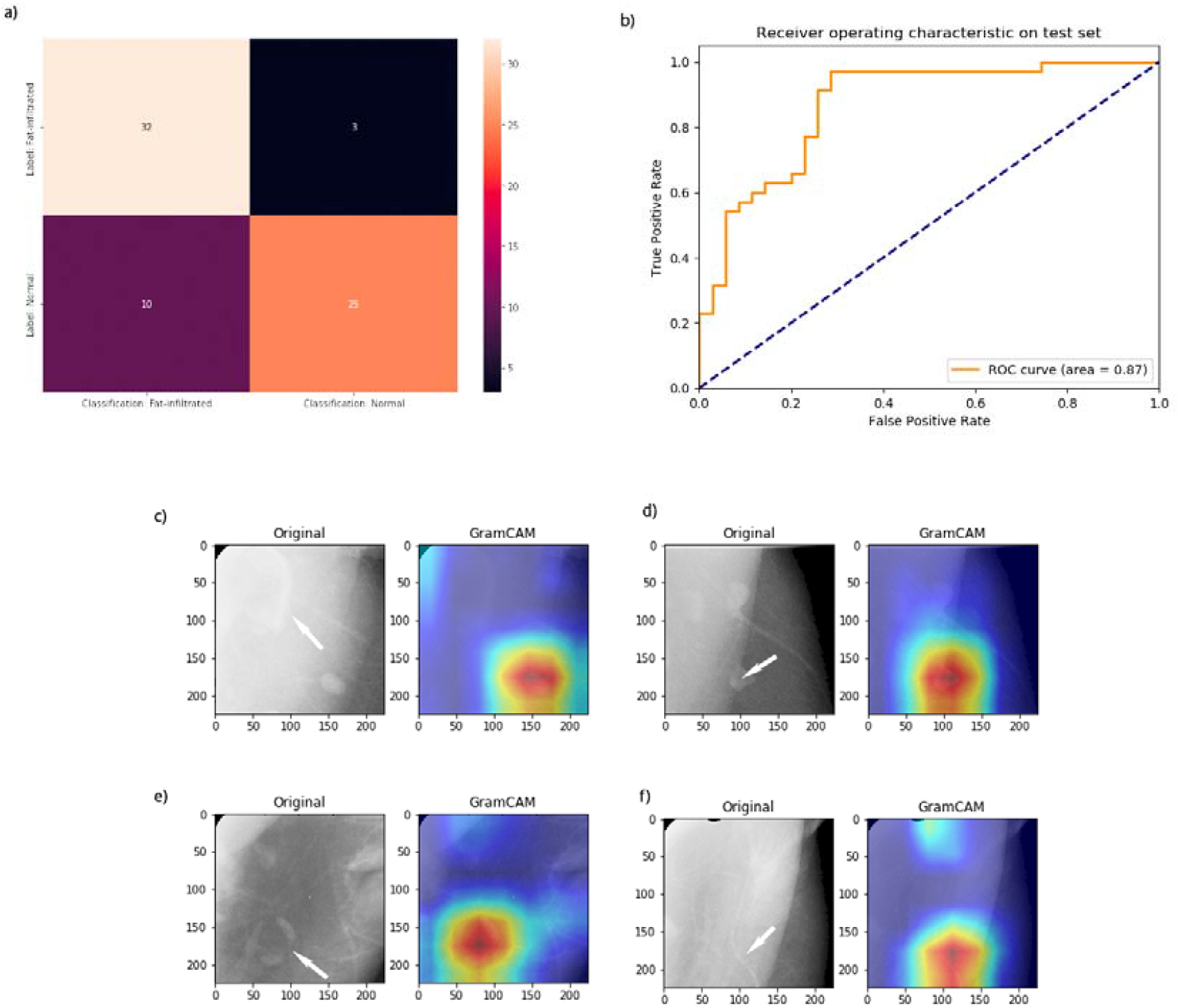
Visualization of model performance and error analysis on the external DDSM dataset. a) Confusion matrix of model classification. b) ROC curve of model prediction. c) A false negative classification resulted from misidentifying the fat-infiltrated LN (arrow-pointed) due to a low contrast from the background and focusing only on the normal node. d) A false-positive case possibly related to the overlying pectoralis muscle and a small node with a lucent center (arrow-pointed). The highlighted node has a lucent center with a normal size relative to the surrounding normal node. e) A false-positive case due to misclassification of multiple normal nodes with space in between as fat infiltrated (arrow-pointed). f) Misclassification of vessels (arrow-pointed) as fat-infiltrated nodes.

**Table 3.** Model Performance on the External Dataset

| *Model* | *Accuracy* | *Precision* | *Sensitivity* | *Specificity* | *F1 Score* | *AU-ROC* |
| --- | --- | --- | --- | --- | --- | --- |
| ***Resnet18***<br>***(Stage 2)*** | **0.82**<br>**(0.77, 0.86)** | **0.76**<br>**(0.69, 0.83)** | **0.92**<br>**(0.87, 0.96)** | **0.72**<br>**(0.64, 0.79)** | **0.81**<br>**(0.78, 0.88)** | **0.87**<br>**(0.82, 0.91)** |

## Discussion

Emerging evidence shows that fat-infiltrated axillary LNs identified in breast imaging studies are associated with obesity and obesity-related pathology. These findings suggest that expanded lymph node hilar fat may represent ectopic fat deposition within the immune-lymphatic system. Large-scale studies are needed to confirm axillary fatty nodes as a potential imaging biomarker of obesity-associated illness among patients. Due to the time-intensive nature of manual lymph node evaluation, a machine learning technique that can automatically identify and classify fatty nodes in large datasets is needed to enable further investigation of the role of ectopic fat deposition within the immune-lymphatic system. In this study, we developed an automated end-to-end DL-based pipeline, from image preprocessing to the classification of the fat-infiltrated axillary LN status, from digital screening mammography without manual annotation of the LNs. Our model achieved an accuracy of 0.97 and a 1.00 AU-ROC in classifying the LN status of an independent internal test dataset. The model’s accuracy and AU-ROC on the external dataset are 0.82 and 0.87, respectively. The model was capable of discriminating LN features from other imaging features with similar radiographic density, including breast tissue, pectoralis muscle, skin folds, and vessels. The model benefited from fine-tuning on oversampled training patches from regions containing these lookalike features. Therefore, this automated approach can categorize the status of fat-infiltrated LNs in digital mammograms in bulk with minimal manual intervention and provide a critically needed tool for further research of fat-infiltrated LNs and their association with obesity-related diseases.

Screening mammography is widely used as a primary tool for early breast cancer detection and has a high utilization among age-eligible females in the United States^28^. Digital imaging studies contain information that is unrelated to the study indication but may be used for patient risk stratification and prediction of future adverse health events. Consideration of these incidental imaging features is referred to as opportunistic imaging. Recent studies have revealed that the added value of opportunistic imaging can improve disease screening and benefit patient care^29^. For example, studies have reported that breast arterial calcification (BAC) detected by mammography screening could be a risk marker for coronary artery disease^30–32^. As a result, many computational approaches have been developed to automatically detect BACs from screening mammography^33,34^. Our automated approach for classifying fat-infiltrated LNs is needed to perform additional studies and determine whether fatty nodes may similarly add extra value to mammography screening as a risk marker for obesity-related disorders.

To date, only a few modest studies have looked at fat-infiltrated LNs visualized on screening mammography. There remains a need for a rapid and generalizable tool to assess the status of fat-infiltrated axillary LNs for future studies. To our knowledge, our study is the first to analyze fat-infiltrated axillary LNs using an automated DL approach. In previous studies, human-extracted features, including expanded hilar fat morphology, hilar and cortical dimensions, and overall LN size, were manually assessed by multiple radiologists^9,11,14^. The measurement and assessment of fat-infiltrated LNs are not readily available in any existing dataset and are resource intensive and time-consuming to collect in bulk. Our end-to-end DL approach has the advantage of automatically learning and extracting radiomic feature representations related to fat-infiltrated LNs directly from mammography images and providing consistent and accurate outcomes on new images. Our model bypasses the manual feature engineering step and requires minimal preprocessing steps, thereby significantly reducing individual bias and the time needed to measure and evaluate the LN status on images. Machine-extracted LN radiomic features can capture objective and detailed morphological and textual characteristics that may not be detectable by visual analysis. These radiomic features can aid image interpretation and provide insight into fat-infiltrated LNs and their disease association in future studies^35,36^. Our model successfully captured the features relevant to the axillary LNs in almost all cases that were verified by GradCAM visualizations, and generated the proper classification on our test sets.

The high sensitivity of 0.92 of our model on the subset of the external CBIS-DDSM dataset confirmed the model’s utility in classifying fat-infiltrated LNs on mammograms from an unseen data distribution. There are several possible explanations for the reduced overall model performance on the external data compared to the internal dataset. DDSM is a collection of digitized analog film-screen mammograms first released in 1997. Due to the inherently low contrast resolution of film screen mammography compared to digital mammography, axillary LNs were more challenging to characterize on the external dataset^37^. Second, the CBIS-DDSM data contain no pixel spacing information and therefore did not allow for an LN measurement as a feature of fat-infiltrated LNs. The labeling of the external dataset relied on the LN morphology without consideration of the LN size and was therefore less informative and more error-prone than the internal dataset. However, our model performed well in characterizing axillary LNs as fatty on the consensus labels of the external dataset using only morphologic criteria. Nonetheless, CBIS-DDSM is currently the only public dataset with a sufficient number of digitized full-field mammograms for external validation. Our model should be re-evaluated when additional digital mammography datasets become publicly available.

The results of this study must be considered with a few limitations. First, all mammography exams used for model development were obtained from a single medical center. Our pipeline with a fixed-sized crop of the axilla worked well in capturing the axillary LNs within a uniform image dataset; however, due to variations in scanning procedures and equipment, the same setting may not work for mammograms acquired from other institutions. Further training and validation of our model on larger datasets across multiple institutions would improve the generalizability of our approach. Therefore, we plan to further validate our model on additional data from external collaborators. Second, our model coarsely localized LNs based on image-level labels and still generated false-positive classifications in cases where multiple normal LNs overlapped and formed a cluster around subcutaneous fat. The model may benefit from segmentation approaches, such as U-Net^38^, to identify individual LNs if detailed pixel-level annotations become available. Last, our model was designed to classify the binary status of LNs as normal versus fat-infiltrated, and the size cutoff between the two classes was selected based on the evidence and previous findings in this domain^9^. In future work, we plan to expand the model to provide a more fine-grained classification of fat-infiltrated LNs, including the degree of hilar fat expansion and the number of fat-expanded LNs. We also plan to extend our study to examine the potential associations between fat-infiltrated LN-related features and breast cancer outcomes, and evaluate the model’s viability as a potential risk stratification tool for obesity and other obesity-related diseases. In summary, our study demonstrated a promising performance of a DL model developed to classify mammographically visualized axillary lymph nodes. The model achieved a high performance on both internal and external test sets. This approach can enable further investigation of the role of fat-infiltrated axillary nodes in obesity-associated deaths.

## Data Availability

A de-identified version of the data and the source code for this analysis can be shared with interested researchers upon reasonable request.

