## Supplementary for "Automated Classification of Fat-infiltrated Axillary Lymph Nodes on Screening Mammograms"

### Supplemental Materials

#### S1: Details of Image Preprocessing and Augmentation.

#### S2: Final Hyperparameters for Each CNN Model.

#### S3: ROC Comparison of Stage 1 Models

#### S4: Visualization of Incorrect Predictions.

### Supplementary S1: Details of Image Preprocessing and Augmentation

The input mammograms were received with text markers on the background that were unrelated to the target labels. We removed the image background by finding the largest contour of the breast and masking out areas outside of the contour. The contour finding algorithm was developed by Suzuki and Abe and was implemented in OpenCV-Python.^1^ We randomly augmented the images before feeding them into the training process. All images were horizontally or vertically flipped or 90-degree rotated at random. Image sharpening, contrast addition, salt and pepper and Gaussian noise additions were performed with a probability of 0.3 for every image. The image augmentation was implemented using the *Imgaug* Python library.^2^

### Supplementary S2

### Table 1. Final Hyperparameters for Each CNN Model.

| **Model** | **Batch Size** | **Initial Learning Rate** | **Epochs Trained** |
| --- | --- | --- | --- |
| Resnet18 | 8 | $1\times{10}^{-4}$ | 30 |
| VGG16 | 8 | $2.5\times{10}^{-5}$ | 14 |
| Densenet121 | 16 | $1\times{10}^{-4}$ | 20 |

### Supplementary S3: ROC Comparison of Stage 1 Models

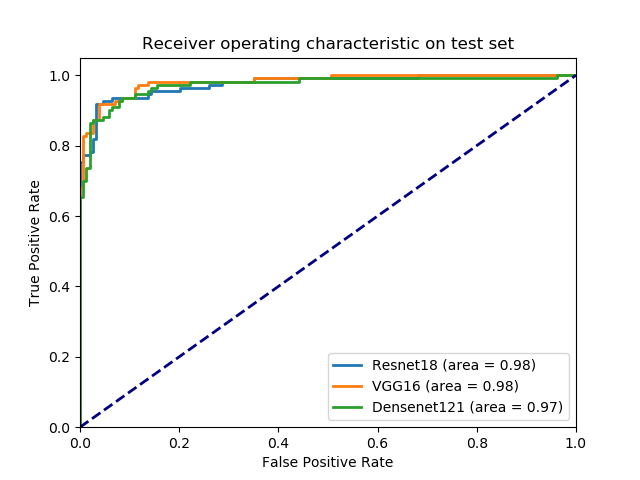

**Figure 1. ROC of the Stage 1 models on the internal test set of axillary patches.** The ResNet18 model achieved an AUC of 0.98 (95% CI: 0.96-0.99) and was used as the final model, while VGG16 achieved an AUC of 0.98 (95% CI: 0.97-0.99), and Densenet121 achieved an AUC of 0.97 (95% CI: 0.95-0.99).

### Supplementary S4: Visualization of Incorrect Predictions

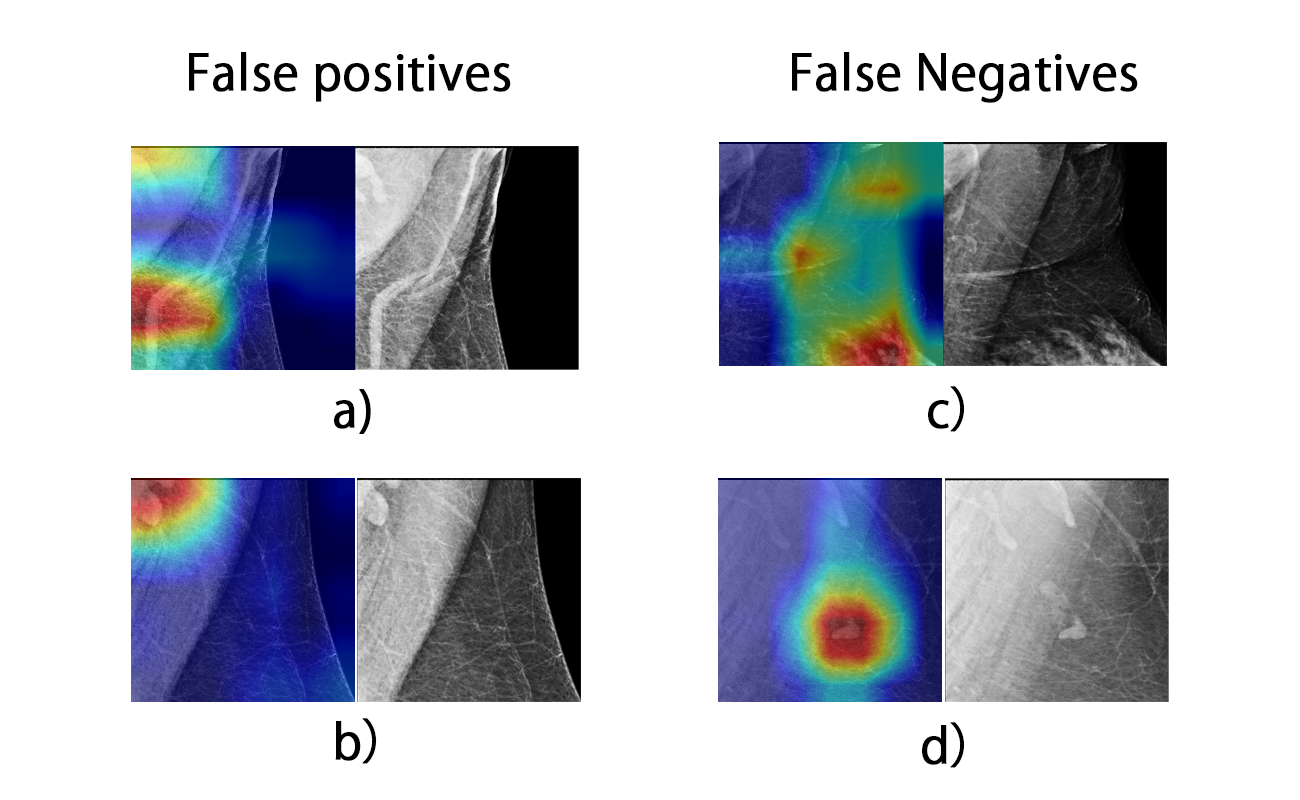

**Figure 2.** **Example of a Grad-CAM visualization of incorrect predictions by Stage 1 Resnet18 model.** The model incorrectly highlighted **a)** scar tissue and **b)** multiple normal LNs as fat-infiltrated. **c)** The model missed fatty LNs and highlighted breast tissue as normal LNs. **d)** The model only highlighted the normal LNs where a mixture of fatty and normal LNs existed.
